# Reengineering a machine learning phenotype to adapt to the changing COVID-19 landscape: A study from the N3C and RECOVER consortia

**DOI:** 10.1101/2023.12.08.23299718

**Authors:** Miles Crosskey, Tomas McIntee, Sandy Preiss, Daniel Brannock, Yun Jae Yoo, Emily Hadley, Frank Blancero, Rob Chew, Johanna Loomba, Abhishek Bhatia, Christopher G. Chute, Melissa Haendel, Richard Moffitt, Emily Pfaff, N3C Consortium, the RECOVER EHR Cohort

## Abstract

**Background:** In 2021, we used the National COVID Cohort Collaborative (N3C) as part of the NIH RECOVER Initiative to develop a machine learning (ML) pipeline to identify patients with a high probability of having post-acute sequelae of SARS-CoV-2 infection (PASC), or Long COVID. However, the increased home testing, missing documentation, and reinfections that characterize the latter years of the pandemic necessitate reengineering our original model to account for these changes in the COVID-19 research landscape.

**Methods:** Our updated XGBoost model gathers data for each patient in overlapping 100-day periods that progress through time, and issues a probability of Long COVID for each 100-day period. If a patient has known acute COVID-19 during any 100-day window (including reinfections), we censor the data from 7 days prior to the diagnosis/positive test date through 28 days after. These fixed time windows replace the prior model’s reliance on a documented COVID-19 index date to anchor its data collection, and are able to account for reinfections.

**Results:** The updated model achieves an area under the receiver operating characteristic curve of 0.90. Precision and recall can be adjusted according to a given use case, depending on whether greater sensitivity or specificity is warranted.

**Discussion:** By eschewing the COVID-19 index date as an anchor point for analysis, we are now able to assess the probability of Long COVID among patients who may have tested at home, or with suspected (but untested) cases of COVID-19, or multiple SARS-CoV-2 reinfections. We view this exercise as a model for maintaining and updating any ML pipeline used for clinical research and operations.

## BACKGROUND

The electronic health record (EHR) is a rich source of data to study post-acute sequelae of SARS-CoV-2 infection (PASC), or Long COVID, resulting in findings around common symptoms,^1^ risk factors,^2^ subphenotypes,^3,4^ and longitudinal trajectories.^5^ However, there are challenges in using the EHR to study this condition, including missing data, idiosyncratic coding practices, and selection bias.^6^ These challenges should not prevent us from using EHRs as a tool for Long COVID research, but does require thoughtfulness about methods and caveats to ensure utility, rigor, and reproducibility.

Much has changed as we enter the fifth year of COVID-19. Widely available home testing means that EHRs are missing many cases; the CDC’s mandate to collect and report national COVID-19 case counts has ended;^7^ and a high number of Americans with at least one (and often multiple) SARS-CoV-2 infections causes difficulty in identifying control populations within clinical data.^8^ For these reasons, we cannot rely on the same analytical methods that worked earlier in the pandemic to research either COVID-19 or Long COVID, and instead must adapt to the changing landscape.

In 2021, we used the National COVID Cohort Collaborative (N3C)^9^ as part of the NIH RECOVER Initiative^10^ to develop a machine learning (ML) pipeline to identify patients with a high probability of having Long COVID.^11^ The ML model’s purpose is to use information from EHR data to predict missing Long COVID labels, thus serving as a computable phenotype for Long COVID. The model was performant and generalizable,^12^ but heavily relies on the existence and timing of an index date for a patient’s acute COVID-19 infection. Moreover, the model only considers a patient’s first SARS-CoV-2 infection, as at the time we did not anticipate the now-common occurrence of SARS-CoV-2 reinfections.^13^

Here we describe the significant reengineering of our existing machine learning (ML)-driven Long COVID phenotype to be more inclusive and attuned to more recent years of the COVID-19 pandemic. By eschewing the COVID-19 index date as an anchor point for analysis, we are now able to assess the probability of Long COVID among patients who may have tested at home, or with suspected (but untested) cases of COVID-19, or multiple SARS-CoV-2 reinfections. We view this exercise as a model for maintaining and updating any ML pipeline used for clinical research and operations–as clinical knowledge and circumstances change over time, so too must our methods, even when major reengineering is required.

## METHODS

### Training Data Selection

Accurate identification of Long COVID cases in EHRs continues to be challenging, which spurred us to create our original Long COVID ML model^11^ in 2021 (hereafter referred to as LCM 1.0). In our original model, lacking a gold standard to use as a training set, we defined a “silver standard” of patients who had visited a Long COVID specialty clinic at least once. Shortly after the U09.9 ICD-10-CM code was made available for clinical use in October 2021, we updated our silver standard to include patients who had at least one U09.9 code in their EHR. This increased the size of our training set, but inconsistent and idiosyncratic use of that code in practice^6^ likely introduced some amount of noise.

For our new model (LCM 2.0), we leveraged the increased amount of available longitudinal data to now require *two* or more U09.9 codes or *two* or more Long COVID specialty clinic visits to qualify for the training set. This more stringent criterion decreases the risk that a patient is labeled with the Long COVID code due to error or rule-out. All patients included in the training set are 18 years of age or older at the time of their earliest U09.9 diagnosis or clinic visit. The training set contains 36,238 patients with a Long COVID label (per the aforementioned definition), and 36,507 unlabeled patients randomly selected from the population described in Running the Model, below. The validation set contains 1,890 patients with a Long COVID label and 1,899 without.

### Time Windows

LCM 1.0 used each patient’s first acute COVID-19 date as an anchor point, basing its predictions on data sourced from a set number of days before and after that time. As LCM 2.0 does not require acute COVID-19 date(s), we developed a new anchoring method using set time windows that apply to all patients–not just those with COVID-19 index dates. LCM 2.0 gathers data for each patient in overlapping 100-day periods that progress through time, and issues a probability of Long COVID for each 100-day period. If a patient has known acute COVID-19 during any 100-day window (including reinfections), we censor the data from 7 days prior to the diagnosis/positive test date through 28 days after. This prevents the model from confusing the symptoms of acute COVID-19 for Long COVID. A comparison between the anchoring methods of LCM 1.0 and 2.0 is illustrated in **Figure 1**.

**Figure 1.**
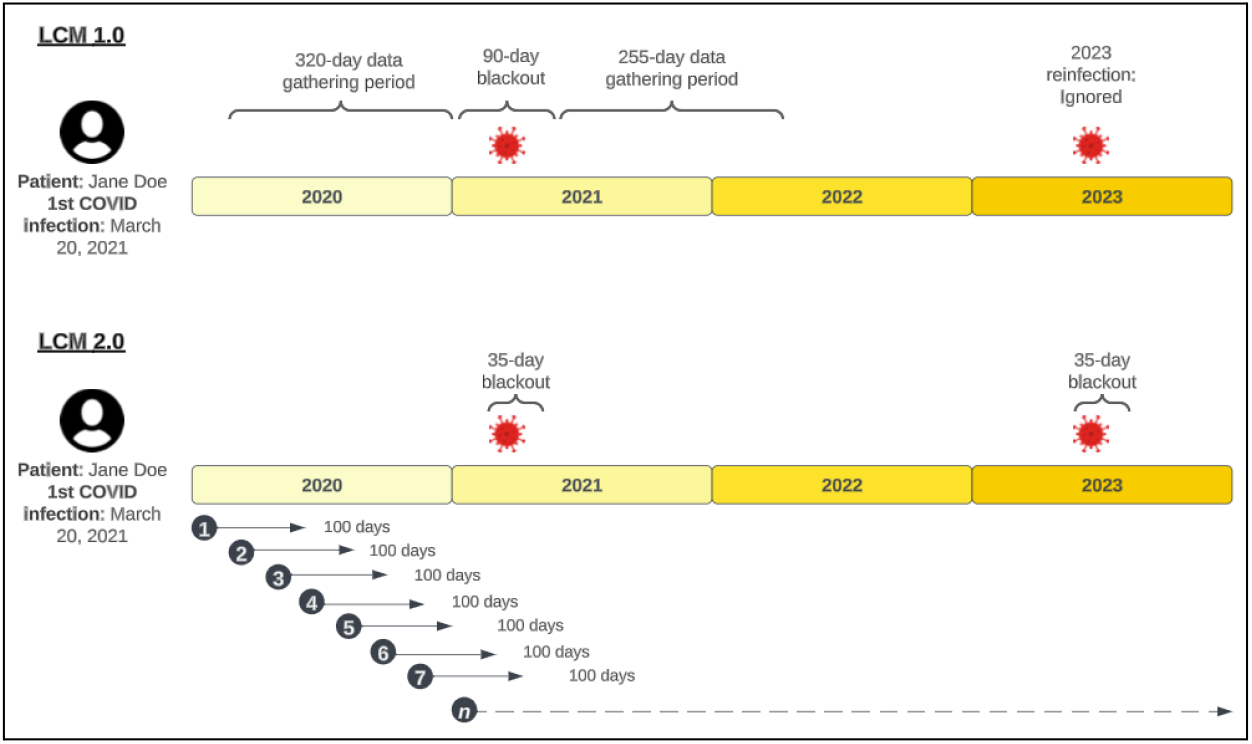
Model “anchor” methods. LCM 1.0 (top) used the patient’s first acute COVID-19 instance to determine the time window in which it gathered evidence for its prediction. The 90 days surrounding the acute infection (45 days before, 45 days after) were censored from data gathering. Reinfections were not taken into account. In LCM 2.0, all patients are assigned probabilities for set windows, where only data from those 100-day windows are considered. Each 100-day window overlaps the prior window by 70 days.The 35 days surrounding any acute infection (7 days before, 28 days after) are censored from data gathering.

### Feature Selection

N3C standardizes on the OMOP clinical data model, which uses SNOMED as a standard vocabulary for diagnoses. However, using every unique SNOMED concept as a model feature would (1) result in thousands of features and major computational inefficiency and (2) be so granular as to potentially obscure important patterns. For this reason, we opted to “roll up” more granular SNOMED concepts to parent concepts, cutting down the initial number of possible diagnosis features from 50,349 (the number of unique codes across the population described in Running the Model, below) to 9,623. As SNOMED is multi-hierarchical (i.e., a child concept can have many parents) and the depth of its leaf nodes is highly variable, we needed to develop an algorithm to select optimal parent concepts using consistent logic. This roll-up method is described in **Supplemental Methods**.

To improve efficiency and avoid over-fitting, we prune this list of features further by assessing the covariance of each selected parent concept with the label from the training set–”1” for Long COVID, “0” for not Long COVID. We select the top 200 concepts with the highest covariance to serve as the model’s diagnosis features. We purposely exclude three features that are essentially COVID-19 and Long COVID labels (e.g., the SNOMED equivalents of the ICD-10 codes U07.1 [COVID-19], U09.9 [Post-COVID condition] and B94.8 [Sequelae of infectious disease, used as a proxy for U09.9 prior to its release]).

Additional features include patient age, number of unique outpatient days, number of unique inpatient days, and patient sex. Continuous variables are binned to reduce overfitting–age is binned into 10-year bins, while the number of visits is binned into [0-2, 3-10, 11-inf).

### Running the Model

The model was constructed using the XGBoost Python package. Once trained, we ran the model on a subset of N3C patients (*n* = 5,875,065) that have at least one of the following at any time between 1/1/2020 and 06/22/2023: (1) a U07.1 (COVID-19) diagnosis code, (2) a positive SARS-CoV-2 test, (3) a U09.9 (PASC) diagnosis code, (4) a prescription for Paxlovid or Remdesivir, or (5) an M35.81 (MIS-C) diagnosis code. While this is a cohort highly enriched with COVID-19 cases, a COVID-19 index date is not required for the model, and is only used to identify blackout dates if it exists. Each patient is given a model score that predicts PASC status for each 100-day window in which they are >= 18 years of age.

### Threshold Selection

Choosing an appropriate threshold is critical for effective utilization of algorithms, particularly when the objective is to obtain a binary decision rather than a continuous confidence score. The threshold serves as the mechanism through which false positive errors are weighed against false negative errors.

Several approaches for selecting a threshold are available, and there is no single method that is generalizable for all use cases.^14^ One common way to select a threshold is to maximize the Youden Index, which maximizes the true positive rate minus the false positive rate–the percent correct (PC). This analysis is done on the validation set. For LCM 2.0, the maximization of PC is achieved at a threshold of 0.9, resulting in a false positive rate of 0.16 and a true positive rate of 0.83 (**Figure 2**).

**Figure 2.**
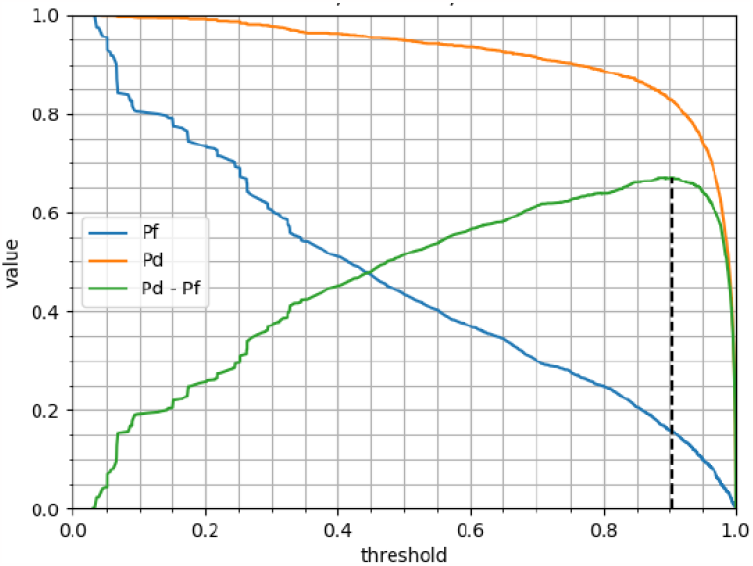
Threshold Selection. Three curves are plotted on this chart. The x-axis represents the threshold value. The lines correspond to different quantities used to measure performance. The blue curve (Pf) is the probability of false alarm. The orange curve (Pd) is the probability of detection, as determined by the validation set. We set the threshold (0.9) to maximize the Youden Index (Pd - Pf), shown in green.

### Historical Controls and False Alarm Rates

Long COVID has many overlapping symptoms with other conditions such as myalgic encephalomyelitis/chronic fatigue syndrome (ME/CFS),^15^ other post-viral syndromes (such as sequelae triggered by influenza),^16^ dysautonomia,^17,18^ and various respiratory illnesses. In many cases, the data available to the model is not detailed enough to distinguish between these conditions and Long COVID. We also know that there are likely a number of undiagnosed patients in the training set who have Long COVID, making it difficult to measure a true false positive rate.

To better understand the false positive rate, we leveraged the same RECOVER analysis cohort described in Running the Model, but swapped in the cohort’s EHR data from 2018-2019, prior to the existence of COVID-19. 5,572,579 patients in the RECOVER analysis cohort had data available for this purpose and met the minimum age criterion during the earlier time period. We compared the distribution of model scores for 2018-2019 to those of 2020-2023 (shown in **Figure 3**), theorizing that the magnitude of patients with high scores in 2019 could serve as a rough estimate of the false positive rate in the COVID period. This is a richer source of evidence of false positivity than the default option of declaring all patients with high scores but no Long COVID label as false positives. In reality, unlabelled patients may be false negatives with undiagnosed Long COVID–a known challenge in assessing performance of diagnostic machine learning models.^19^

**Figure 3.**
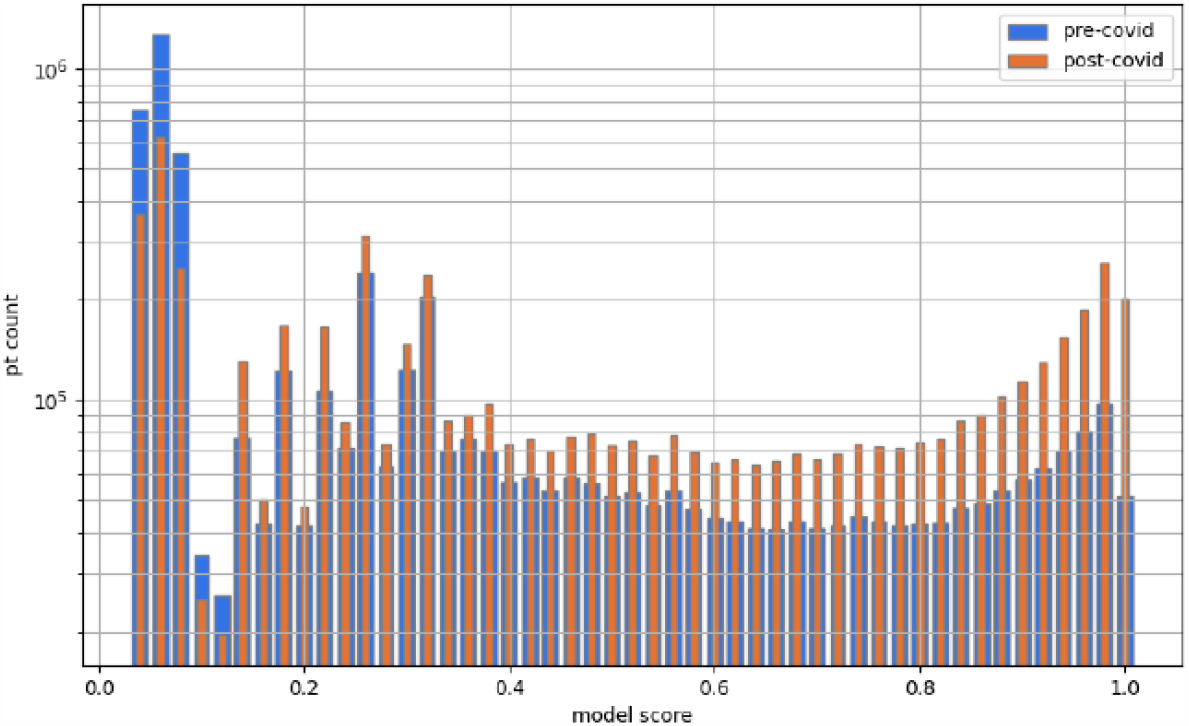
Histogram of control study. Here we show the distribution of model scores for the same population using (1) 2019 data, before COVID-19 (blue) and (2) 2020-2023 data (orange). High model scores in blue represent those patients who look like Long COVID patients, but are in fact false positives, as COVID-19 did not yet exist. We notice a significant shift toward higher model scores in the COVID-19 period, but still a substantial amount of false alarms at various thresholds. Note that counts on the y-axis are on a log scale.

## RESULTS

### Distribution of Model Scores

The distribution of model scores across the full RECOVER cohort (n=5,875,065) is shown in **Figure 4**.

**Figure 4.**
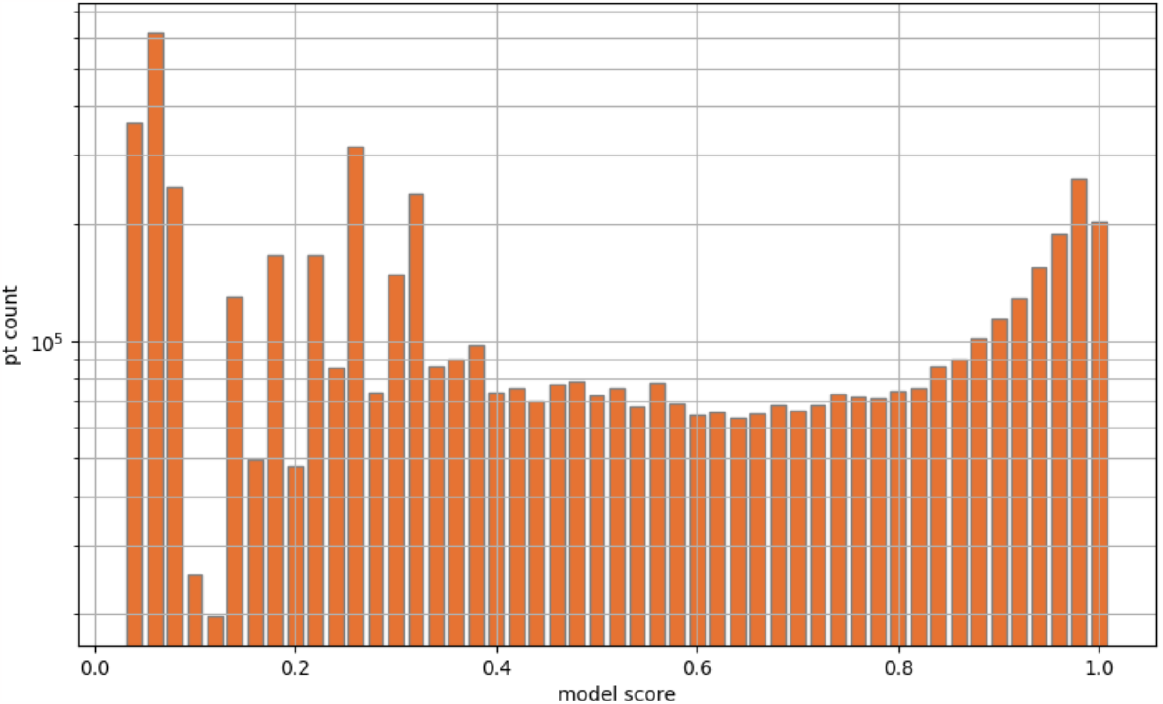
Distribution of model scores. When the model is run over the full RECOVER cohort (n = 5,875,065), predicted probabilities of Long COVID are distributed as shown. Using our threshold of 0.9, 1,051,045 patients are labeled as probable Long COVID. Note that counts on the y-axis are on a log scale.

This distribution of model scores gives us a Long COVID prevalence of 17.9% in this COVID-enriched population, using the 0.9 threshold. However, using the historical control experiment detailed in Methods, we can estimate the proportion of these cases that may be false positives, and thus change our prevalence estimate. Using the 0.9 threshold with the historical controls, we calculate a false positive rate of 7.5%–the difference between 0, which should be the Long COVID prevalence in the pre-COVID era, and 7.5%, the proportion of historical control patients with a model score >= 0.9. Subtracting from 17.9% results in an estimated prevalence of 10.4%.

### Model Performance

Our goal is not only to determine whether a patient is likely to have Long COVID, but also to determine whether a patient is likely to have Long COVID *in a specific time window*. We define the “correct” window for a labeled patient as the latest window that contains a patient’s first U09.9 code or Long COVID clinic visit. We randomly select “correct” windows for unlabeled patients. We consider the model to be correct when a labeled patient has a score above our 0.9 threshold in this window, or an unlabeled patient has a score below the threshold. **Figure 5** shows the receiver operating characteristic (ROC) curve for the model calculated on both a person basis and a window basis.

**Figure 5.**
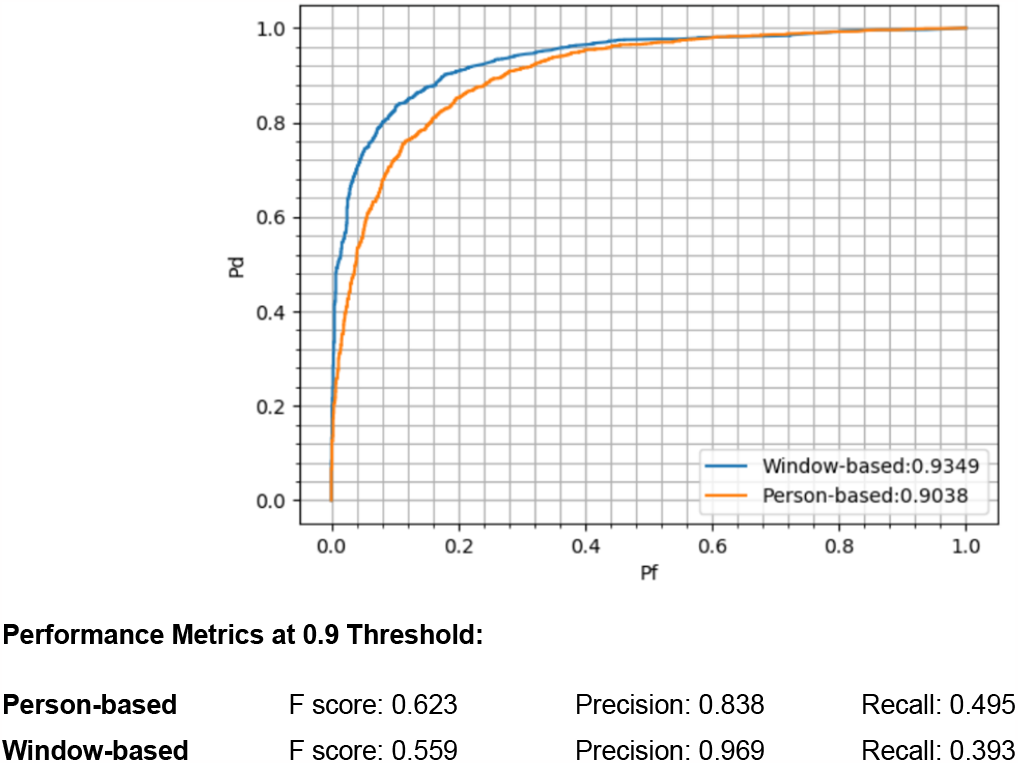
Model performance. Model performance can be measured in two ways, visualized on the receiver operating characteristic (ROC) curve. The first is person-based (orange curve), where we use a patient’s maximum model score over all time to classify them (i.e., “has this patient had Long COVID ever?”). The second is window-based (blue curve), where we use the model score for a specific patient-window to classify that window (i.e., “did this patient have Long COVID in this time window?”).

As noted in Threshold Selection above, our 0.9 threshold was selected to purposefully maximize true positives and minimize false positives. For this reason, we sacrifice recall to maximize precision. Alternative thresholds result in different balances between precision and recall, and could be adjusted as desired for a specific use case.

### Important Features

Table 1 lists the top 20 features of the model as determined by each feature’s Shapley (importance) value. The full list of features with Shapley values is available in **Supplemental Results**.

**Table 1.**
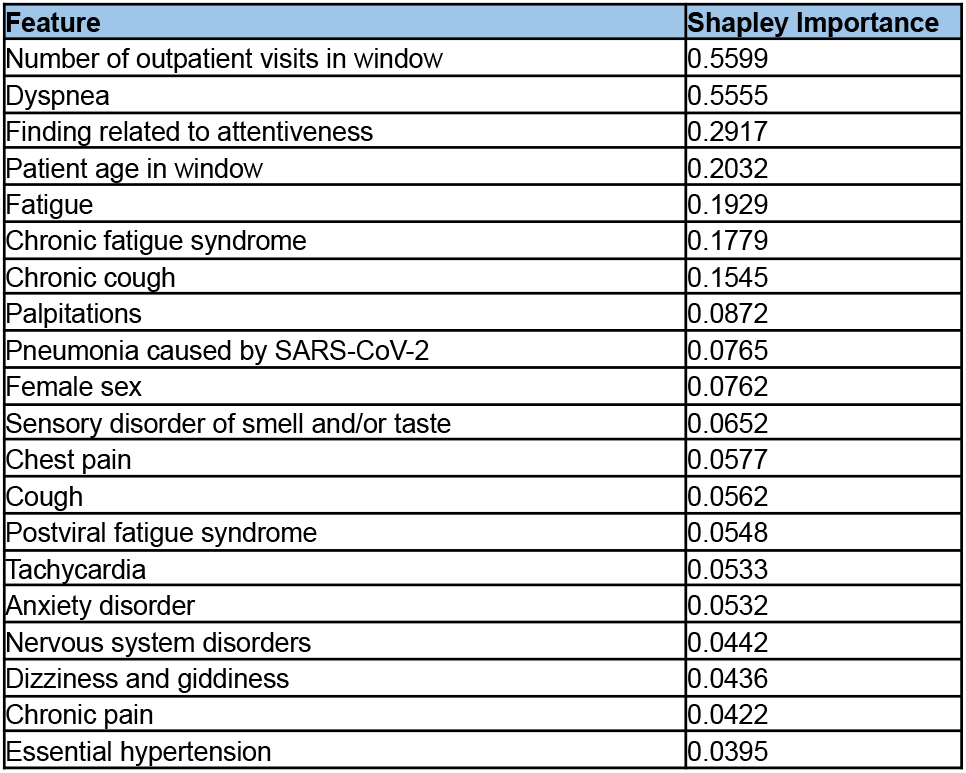
The model’s top 20 features with their Shapley Importance. All of the top 20 features are more likely to increase a given patient’s model score than decrease it when present, other than patient age, which has a nonlinear relationship to the label, and Essential Hypertension, which is more likely to decrease the score.

| Feature | Shapley Importance |
| --- | --- |
| Number of outpatient visits in window | 0.5599 |
| Dyspnea | 0.5555 |
| Finding related to attentiveness | 0.2917 |
| Patient age in window | 0.2032 |
| Fatigue | 0.1929 |
| Chronic fatigue syndrome | 0.1779 |
| Chronic cough | 0.1545 |
| Palpitations | 0.0872 |
| Pneumonia caused by SARS-CoV-2 | 0.0765 |
| Female sex | 0.0762 |
| Sensory disorder of smell and/or taste | 0.0652 |
| Chest pain | 0.0577 |
| Cough | 0.0562 |
| Postviral fatigue syndrome | 0.0548 |
| Tachycardia | 0.0533 |
| Anxiety disorder | 0.0532 |
| Nervous system disorders | 0.0442 |
| Dizziness and giddiness | 0.0436 |
| Chronic pain | 0.0422 |
| Essential hypertension | 0.0395 |

## DISCUSSION

Our updated Long COVID ML computable phenotype reengineers prior work to adapt to the changing landscape of COVID-19. LCM 2.0 performs similarly to its predecessor while (1) removing dependency on date of acute COVID-19 infection as a feature and (2) censoring data from known reinfections.

### Model Explainability

A key test for the generalizability of any ML model is its explainability, or whether the features it relies on the most pass the “common sense” test. Models that rely on unusual features to make decisions may be latching onto idiosyncratic patterns in the training data that would not necessarily translate to external datasets. Our top model features (as shown in **Table 1** and expanded on in **Supplemental Table 1**) include pulmonary, cardiac, and neurologic symptoms that are well-supported by Long COVID literature. Non-symptom-related, continuous variables such as healthcare utilization rates are binned to avoid overfitting and overreliance on patterns unique to the N3C population. As was true of the prior version,^12^ we believe this new model version will be translatable to other sites and consortia with OMOP implementations to promote reuse and reproducibility.

### Inclusivity and the Risk of False Positives

As home COVID-19 testing is (and will continue to be) extremely commonplace, we felt it critical to reengineer our Long COVID model to work without specific EHR documentation of acute COVID-19. This change also allows us to account for (1) unlabeled COVID patients in early-to-mid 2020, when tests were scarce and the U07.1 diagnosis code had not yet been released, and (2) patients who have had COVID but did not seek care for it. In general, this inclusivity is desirable–leaving these populations out of data-driven COVID-19 and Long COVID research may result in selection bias toward patients who are sicker in general, have more access to specialty care, or had more severe COVID-19.

However, with increased sensitivity comes a higher risk of false positives. Our 0.9 threshold was selected to minimize these false positives, though at the expense of high recall. Regardless, as shown in our historical control experiment (**Figure 3)**, there is clearly a population of patients who match the Long COVID symptom profile but do not have Long COVID. Long COVID symptoms are numerous and affect multiple body systems, leading to unavoidable overlap with many other diseases. For this reason, downstream researchers using these model results may wish to consider imposing additional inclusion criteria as desired (e.g., to require documentation of a positive SARS-CoV-2 test in addition to a model score above the threshold), particularly for research use cases requiring higher specificity. Applying additional criteria downstream of the model, as opposed to prior to running the model, enables maximal flexibility.

### Limitations

With the lack of a consensus clinical definition of Long COVID,^20^ assessment of the true accuracy of algorithms like ours is challenging. Moreover, the lack of a consistent estimate for Long COVID’s prevalence^21^ makes it difficult to determine an appropriate model threshold using real-world evidence. Assessing performance using the U09.9 label as ground truth is the most readily available option, but the inconsistency of the code’s use by providers and its late availability^4^ mean that many patients with Long COVID lack the label. As with any ML phenotype, model output should be used thoughtfully, and additional selection criteria should be imposed for use cases where specificity is paramount.

Many Long COVID symptoms (e.g., post-exertional malaise, brain fog) are not well-represented in the EHR, either because they do not have a discrete ICD-10-CM code or are more commonly referenced in free-text clinical notes than structured data. These features thus do not have the opportunity to contribute to the model–however, that should not be interpreted to mean that these symptoms are unreported or not experienced by this patient population.

## CONCLUSION

As ML is increasingly used as a computable phenotyping tool, it is incumbent upon its users to ensure that models are not only created, but maintained and updated over time. As an example of this principle, the landscape of COVID-19 and Long COVID has changed significantly over the course of the pandemic, and so too must our model. We reengineered our ML Long COVID phenotype to respond to these changes, solving some challenges from the prior model (not accounting for reinfection, requiring an acute COVID-19 index date) and encountering new ones (increased risk of false positives). We believe this new model to be generalizable for use as a foundation of data-driven Long COVID research.

## Data Availability

The N3C data transfer to NCATS is performed under a Johns Hopkins University reliance protocol (IRB00249128) or individual site agreements with the NIH. The N3C Data Enclave is managed under the authority of the NIH; more information can be found at ncats.nih.gov/n3c/resources. Enclave data is protected, and can be accessed for COVID-19-related research with an institutional review board-approved protocol and data use request. The Data Use Request ID for this study is RP-5677B5. Enclave and data access instructions can be found at https://covid.cd2h.org/for-researchers. All code used to produce the analyses in this manuscript is available within the N3C Data Enclave to users with valid login credentials to support reproducibility.

## AUTHOR CONTRIBUTIONS

Authorship has been determined according to ICMJE recommendations.

## ACKNOWLEDGEMENTS

The analyses described in this manuscript were conducted with data or tools accessed through the NCATS N3C Data Enclave https://covid.cd2h.org and N3C Attribution & Publication Policy v 1.2-2020-08-25b supported by NCATS U24 TR002306, Axle Informatics Subcontract: NCATS-P00438-B, and by the RECOVER Initiative (OT2HL161847–01). The content is solely the responsibility of the authors and does not necessarily represent the official views of the RECOVER Program, the NIH or other funders. This research was possible because of the patients whose information is included within the data and the organizations (https://ncats.nih.gov/n3c/resources/data-contribution/data-transfer-agreement-signatories) and scientists who have contributed to the on-going development of this community resource [https://doi.org/10.1093/jamia/ocaa196]. We would also like to thank the National Community Engagement Group (NCEG), all patient, caregiver and community Representatives, and all the participants enrolled in the RECOVER Initiative.

We also acknowledge the following institutions whose data is released or pending:

## Available

Advocate Health Care Network — UL1TR002389: The Institute for Translational Medicine (ITM) • Aurora Health Care Inc — UL1TR002373: Wisconsin Network For Health Research • Boston University Medical Campus — UL1TR001430: Boston University Clinical and Translational Science Institute • Brown University — U54GM115677: Advance Clinical Translational Research (Advance-CTR) • Carilion Clinic — UL1TR003015: iTHRIV Integrated Translational health Research Institute of Virginia • Case Western Reserve University — UL1TR002548: The Clinical & Translational Science Collaborative of Cleveland (CTSC) • Charleston Area Medical Center — U54GM104942: West Virginia Clinical and Translational Science Institute (WVCTSI) • Children’s Hospital Colorado — UL1TR002535: Colorado Clinical and Translational Sciences Institute • Columbia University Irving Medical Center — UL1TR001873: Irving Institute for Clinical and Translational Research • Dartmouth College — None (Voluntary) Duke University — UL1TR002553: Duke Clinical and Translational Science Institute • George Washington Children’s Research Institute — UL1TR001876: Clinical and Translational Science Institute at Children’s National (CTSA-CN) • George Washington University — UL1TR001876: Clinical and Translational Science Institute at Children’s National (CTSA-CN) • Harvard Medical School — UL1TR002541: Harvard Catalyst • Indiana University School of Medicine — UL1TR002529: Indiana Clinical and Translational Science Institute • Johns Hopkins University — UL1TR003098: Johns Hopkins Institute for Clinical and Translational Research • Louisiana Public Health Institute — None (Voluntary) • Loyola Medicine Loyola University Medical Center • Loyola University Medical Center — UL1TR002389: The Institute for Translational Medicine (ITM) • Maine Medical Center — U54GM115516: Northern New England Clinical & Translational Research (NNE-CTR) Network • Mary Hitchcock Memorial Hospital & Dartmouth Hitchcock Clinic — None (Voluntary) • Massachusetts General Brigham UL1TR002541: Harvard Catalyst • Mayo Clinic Rochester — UL1TR002377: Mayo Clinic Center for Clinical and Translational Science (CCaTS) • Medical University of South Carolina — UL1TR001450: South Carolina Clinical & Translational Research Institute (SCTR) • MITRE Corporation — None (Voluntary) • Montefiore Medical Center — UL1TR002556: Institute for Clinical and Translational Research at Einstein and Montefiore • Nemours — U54GM104941: Delaware CTR ACCEL Program • NorthShore University HealthSystem — UL1TR002389: The Institute for Translational Medicine (ITM) • Northwestern University at Chicago — UL1TR001422: Northwestern University Clinical and Translational Science Institute (NUCATS) • OCHIN — INV-018455: Bill and Melinda Gates Foundation grant to Sage Bionetworks • Oregon Health & Science University — UL1TR002369: Oregon Clinical and Translational Research Institute • Penn State Health Milton S. Hershey Medical Center — UL1TR002014: Penn State Clinical and Translational Science Institute • Rush University Medical Center — UL1TR002389: The Institute for Translational Medicine (ITM) • Rutgers, The State University of New Jersey — UL1TR003017: New Jersey Alliance for Clinical and Translational Science • Stony Brook University — U24TR002306 • The Alliance at the University of Puerto Rico, Medical Sciences Campus — U54GM133807: Hispanic Alliance for Clinical and Translational Research (The Alliance) • The Ohio State University — UL1TR002733: Center for Clinical and Translational Science • The State University of New York at Buffalo — UL1TR001412: Clinical and Translational Science Institute • The University of Chicago — UL1TR002389: The Institute for Translational Medicine (ITM) • The University of Iowa — UL1TR002537: Institute for Clinical and Translational Science • The University of Miami Leonard M. Miller School of Medicine — UL1TR002736: University of Miami Clinical and Translational Science Institute • The University of Michigan at Ann Arbor — UL1TR002240: Michigan Institute for Clinical and Health Research The University of Texas Health Science Center at Houston — UL1TR003167: Center for Clinical and Translational Sciences (CCTS) • The University of Texas Medical Branch at Galveston — UL1TR001439: The Institute for Translational Sciences • The University of Utah — UL1TR002538: Uhealth Center for Clinical and Translational Science • Tufts Medical Center — UL1TR002544: Tufts Clinical and Translational Science Institute • Tulane University — UL1TR003096: Center for Clinical and Translational Science • The Queens Medical Center — None (Voluntary) • University Medical Center New Orleans — U54GM104940: Louisiana Clinical and Translational Science (LA CaTS) Center • University of Alabama at Birmingham — UL1TR003096: Center for Clinical and Translational Science • University of Arkansas for Medical Sciences — UL1TR003107: UAMS Translational Research Institute • University of Cincinnati — UL1TR001425: Center for Clinical and Translational Science and Training • University of Colorado Denver, Anschutz Medical Campus — UL1TR002535: Colorado Clinical and Translational Sciences Institute • University of Illinois at Chicago — UL1TR002003: UIC Center for Clinical and Translational Science • University of Kansas Medical Center — UL1TR002366: Frontiers: University of Kansas Clinical and Translational Science Institute • University of Kentucky — UL1TR001998: UK Center for Clinical and Translational Science • University of Massachusetts Medical School Worcester — UL1TR001453: The UMass Center for Clinical and Translational Science (UMCCTS) • University Medical Center of Southern Nevada — None (voluntary) • University of Minnesota — UL1TR002494: Clinical and Translational Science Institute • University of Mississippi Medical Center — U54GM115428: Mississippi Center for Clinical and Translational Research (CCTR) • University of Nebraska Medical Center — U54GM115458: Great Plains IDeA-Clinical & Translational Research • University of North Carolina at Chapel Hill — UL1TR002489: North Carolina Translational and Clinical Science Institute • University of Oklahoma Health Sciences Center — U54GM104938: Oklahoma Clinical and Translational Science Institute (OCTSI) • University of Pittsburgh — UL1TR001857: The Clinical and Translational Science Institute (CTSI) • University of Pennsylvania — UL1TR001878: Institute for Translational Medicine and Therapeutics • University of Rochester — UL1TR002001: UR Clinical & Translational Science Institute • University of Southern California — UL1TR001855: The Southern California Clinical and Translational Science Institute (SC CTSI) • University of Vermont — U54GM115516: Northern New England Clinical & Translational Research (NNE-CTR) Network • University of Virginia — UL1TR003015: iTHRIV Integrated Translational health Research Institute of Virginia • University of Washington — UL1TR002319: Institute of Translational Health Sciences • University of Wisconsin-Madison — UL1TR002373: UW Institute for Clinical and Translational Research • Vanderbilt University Medical Center — UL1TR002243: Vanderbilt Institute for Clinical and Translational Research • Virginia Commonwealth University — UL1TR002649: C. Kenneth and Dianne Wright Center for Clinical and Translational Research • Wake Forest University Health Sciences — UL1TR001420: Wake Forest Clinical and Translational Science Institute • Washington University in St. Louis — UL1TR002345: Institute of Clinical and Translational Sciences • Weill Medical College of Cornell University — UL1TR002384: Weill Cornell Medicine Clinical and Translational Science Center • West Virginia University — U54GM104942: West Virginia Clinical and Translational Science Institute (WVCTSI) Submitted: Icahn School of Medicine at Mount Sinai — UL1TR001433: ConduITS Institute for Translational Sciences • The University of Texas Health Science Center at Tyler — UL1TR003167: Center for Clinical and Translational Sciences (CCTS) • University of California, Davis — UL1TR001860: UCDavis Health Clinical and Translational Science Center • University of California, Irvine — UL1TR001414: The UC Irvine Institute for Clinical and Translational Science (ICTS) • University of California, Los Angeles — UL1TR001881: UCLA Clinical Translational Science Institute • University of California, San Diego — UL1TR001442: Altman Clinical and Translational Research Institute • University of California, San Francisco — UL1TR001872: UCSF Clinical and Translational Science Institute Pending: Arkansas Children’s Hospital — UL1TR003107: UAMS Translational Research Institute • Baylor College of Medicine — None (Voluntary) • Children’s Hospital of Philadelphia — UL1TR001878: Institute for Translational Medicine and Therapeutics • Cincinnati Children’s Hospital Medical Center — UL1TR001425: Center for Clinical and Translational Science and Training • Emory University — UL1TR002378: Georgia Clinical and Translational Science Alliance • HonorHealth — None (Voluntary) • Loyola University Chicago — UL1TR002389: The Institute for Translational Medicine (ITM) • Medical College of Wisconsin — UL1TR001436: Clinical and Translational Science Institute of Southeast Wisconsin • MedStar Health Research Institute — None (Voluntary) • Georgetown University — UL1TR001409: The Georgetown-Howard Universities Center for Clinical and Translational Science (GHUCCTS) • MetroHealth — None (Voluntary) • Montana State University — U54GM115371: American Indian/Alaska Native CTR • NYU Langone Medical Center — UL1TR001445: Langone Health’s Clinical and Translational Science Institute • Ochsner Medical Center — U54GM104940: Louisiana Clinical and Translational Science (LA CaTS) Center • Regenstrief Institute — UL1TR002529: Indiana Clinical and Translational Science Institute • Sanford Research — None (Voluntary) • Stanford University — UL1TR003142: Spectrum: The Stanford Center for Clinical and Translational Research and Education • The Rockefeller University — UL1TR001866: Center for Clinical and Translational Science • The Scripps Research Institute — UL1TR002550: Scripps Research Translational Institute • University of Florida — UL1TR001427: UF Clinical and Translational Science Institute University of New Mexico Health Sciences Center — UL1TR001449: University of New Mexico Clinical and Translational Science Center • University of Texas Health Science Center at San Antonio — UL1TR002645: Institute for Integration of Medicine and Science • Yale New Haven Hospital — UL1TR001863: Yale Center for Clinical Investigation

## SUPPLEMENTAL METHODS

### SNOMED Roll Up

The OMOP concept_ancestor table provides a tabular version of the SNOMED graph and allows us to identify all parent concepts for each SNOMED term, as well as the minimum number of “hops” it takes to get from a child concept to each parent. Joining concept_ancestor to condition_occurrence enables us to determine how many instances of each concept appear in N3C data. Our parent concept selection algorithm uses this information to select optimal roll-ups.

For each feature, we walk up the SNOMED graph (i.e., from child to parent, to grandparent, to great-grandparent) to find all ancestor concepts that have >=25,000 occurrences in the N3C data. Once this set of ancestor concepts are identified for a given child, we select the *closest* ancestor with >=25,000 as the optimal parent.

**Figure S1.**
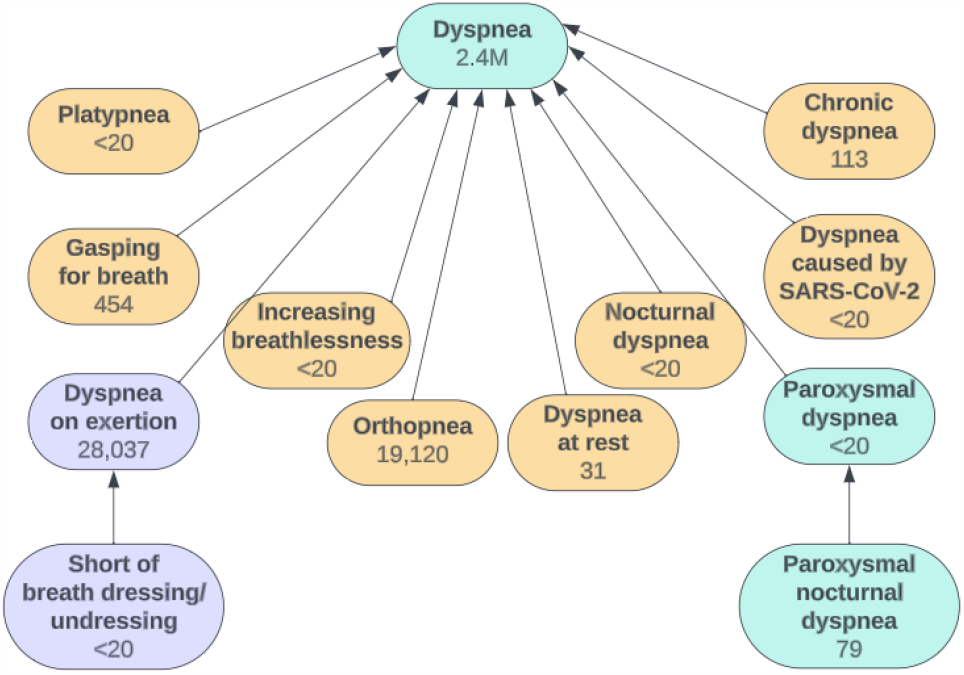
Finding optimal parent concepts. In this example, the two lowermost nodes (Short of breath dressing/undressing and Paroxysmal nocturnal dyspnea) are each rolled up to their optimal parent. The immediate parent of Short of breath dressing/undressing, Dyspnea on exertion, passes the 25,000 instance threshold, making it the optimal parent. For purposes of the model, the few patients with instances of Short of breath dressing/undressing will be relabeled with concept Dyspnea on exertion. The immediate parent of Paroxysmal nocturnal dyspnea, Paroxysmal dyspnea, actually has fewer instances than its child, so we continue walking up the graph. The next parent is Dyspnea, with 2.4 million instances–a much less granular concept, but more optimal as a model feature in this case.

Note that the threshold of 25,000 instances was selected for this use case, in a dataset the size of the N3C data (2,061,673,837 rows in condition_occurrence). This threshold can and should be tailored to the size of the dataset and the condition specificity desired for the given problem.

## SUPPLEMENTAL RESULTS

***Table S1. All model’s features with their Shapley Importance***. *All features are correlated with the Long COVID label, but a negative value in the Direction column signifies that that feature is more likely to decrease a given patient’s model score than increase it*.

[See attached Excel file]

